# Chronic Atrial and Intestinal Dysrhythmia Syndrome: A Distinct Monogenic Cause of Cerebral Small Vessel Disease

**DOI:** 10.64898/2026.08.31.26360967

**Authors:** Caroline Dallaire-Théroux, Ahmad Nehme, Francis Brunet, Christian Berthelot, Marie-Christine Camden, Emilie Bergeron, Mathilde Bizou, Alexandre Dubrac, Philippe Chetaille, Gregor Andelfinger, Steve Verreault

## Abstract

**Objective:** Chronic atrial and intestinal dysrhythmia (CAID) syndrome is a rare autosomal recessive cohesinopathy classically defined by sick sinus syndrome and chronic intestinal pseudo-obstruction; however, emerging evidence suggests an association with cerebral small vessel disease (CSVD). We aimed to characterize the neurological and neuroimaging spectrum of CSVD in CAID syndrome.

**Methods:** We conducted a cross-sectional, retrospective study of 16 French-Canadians with genetically confirmed CAID syndrome. All patients underwent comprehensive neurological assessment. Brain MRI was performed in 14 patients, with CSVD markers evaluated by an expert neuroradiologist according to the STRIVE-2 criteria.

**Results:** The median age at last evaluation was 34 years (range, 19–60); 62.5% were women. Neurological manifestations included migraines (44.4%), mild cerebellar signs (16.7%), and ischemic or hemorrhagic cerebrovascular events (12.5%). MRI showed white matter hyperintensities (92.9%), lacunes (50%) and cerebral microbleeds (85.7%), affecting deep, lobar, and infratentorial regions, with marked cerebellar predominance (11/12; 91.7%); five patients exhibited innumerable microbleeds. Despite the young cohort, moderate-to-severe CSVD was common (median SVD score 1.5, IQR 0–4). Patients with countless microbleeds were older than those with discrete lesions (46.2 vs. 31.2 years; p=0.043). Management of atrial fibrillation required individualized strategies, including left atrial appendage closure, balancing ischemic and hemorrhagic risks.

**Interpretation:** CAID syndrome represents a novel monogenic cause of CSVD, characterized by early, extensive cerebral microbleeds with mixed distribution and distinctive cerebellar predominance. Coexisting congenital cardiac disease and arrhythmias place patients at dual ischemic and hemorrhagic risk. Systematic neurological evaluation and MRI are warranted, particularly prior to antithrombotic therapy.

## INTRODUCTION

Chronic atrial and intestinal dysrhythmia (CAID) syndrome is a rare monogenic cohesinopathy caused by the recessive founder mutation *SGO1 K23E*, encoding the Shugoshin-like 1 protein^1,2^. *SGO1* protects the cohesin complex from separase-mediated cleavage during meiotic cell division. Cultured cells from individuals with homozygous *SGO1* mutations show accelerated cell cycle progression, higher rate of senescence and enhanced TGF-β signaling^1,3^. To date, fewer than 30 affected individuals from approximately 15 families have been reported worldwide^1,4–7^. Most cases involve French-Canadians^1,4^, although patients of White European (Sweden^1^, Belgium^5^) and Asian descent (India^6^, Turkey^7^) have also been described.

Unlike more common cohesinopathies such as Cornelia de Lange syndrome, CAID syndrome is primarily defined by the co-occurrence of sick sinus syndrome (SSS) and chronic intestinal pseudo-obstruction (CIPO), reflecting pacemaker failure in the heart and gastrointestinal tract, presenting within the first four decades of life^1,7,8^. Early descriptions did not report growth or developmental anomalies, nor behavioral or neurological manifestations, apart from a possible association with intracranial aneurysms^9^ and an isolated case with facial dysmorphia and prematurity-related developmental delay^8^. Nevertheless, other congenital abnormalities, particularly cardiac malformations including valvular defects, are frequently observed^2^. As symptoms are absent at birth and emerge after infancy, generally from age five onward, the clinical phenotype is suggested to be associated with aging^3^.

Gastrointestinal manifestations, often preceding cardiac involvement, result from combined neurogenic and myogenic dysfunction, with histopathology demonstrating thinning of smooth muscle layers, disrupted fiber architecture, and fibrosis, implicating defective interstitial cells of Cajal^1,8^. Similarly, sinoatrial node involvement underlies early-onset arrhythmias in homozygous *SGO1* mutation carriers, including chronotropic incompetence, sinoatrial block, and atrial dysrhythmias^1,3^, often requiring permanent pacemaker implantation. Nutritional status, largely dependent on parenteral support and sometimes intestinal transplantation, remains a critical determinant of morbidity and mortality, while device-related complications such as catheter- or electrode-associated septic emboli may further contribute to adverse outcomes^7^.

Although neurological manifestations have historically been considered minimal, emerging evidence suggests a possible association between CAID syndrome and cerebral small vessel disease (CSVD). Young-onset periventricular and pontine white matter hyperintensities (WMH) and multiple cerebral microbleeds (CMB) predominantly affecting the cerebellum have been reported in three patients, raising the hypothesis, among others, that premature vascular cell senescence may contribute to cerebrovascular vulnerability^4,5^. These findings expand the spectrum of CAID syndrome beyond cardiac and gastrointestinal involvement, highlighting the need for comprehensive neurological evaluation and risk stratification in this unique patient population.

This study presents a case series of 16 French-Canadian adult patients with genetically-confirmed CAID syndrome due to *SGO1* mutations, focusing on central nervous system involvement. The primary aim is to characterize the full neurological phenotypic spectrum of this rare disorder, with particular emphasis on CSVD manifestations and their clinical implications.

## METHODS

### Participants

This cross-sectional retrospective study was conducted through a provincial Quebec network collaboration of clinician and researchers with a shared interest and expertise in CAID syndrome. All adults (≥18 years) with genetically confirmed CAID syndrome due to an *SGO1* mutation who were previously enrolled through the Cardiology Service and referred for a neurology consult at the CHU de Québec up to 2025, and who provided informed consent, were included. Eligible participants were identified from an ongoing longitudinal cohort of individuals with CAID syndrome at Hôpital Sainte-Justine and the Centre Hospitalier de l’Université Laval (MP-21-2006-107).

### Clinical Data Collection

All CAID patients underwent comprehensive neurological assessment by a vascular neurologist at Hôpital de l’Enfant-Jésus, CHU de Québec (M.C.C. or S.V.). For this study, data were retrospectively collected from medical records by a local investigator (C.D.T.), up to the last neurological appointment. Cardiology and gastroenterology records were reviewed for additional details. Collected variables included demographics, substance use, vascular risk factors, medical and family history, first CAID presentation, genetic testing, neurological findings, cardiac and gastrointestinal manifestations, related treatments and interventions, antithrombotic exposure, indication for brain magnetic resonance imaging (MRI), and cause of death with postmortem assessment when applicable. Among non-neurological CAID manifestations, SSS was defined as chronic, inappropriate sinus bradycardia with junctional rhythm, sinus pauses and atrial dysrhythmias (frequent atrial premature complexes, atrial tachycardia, flutter, or fibrillation), while CIPO was diagnosed based on mechanical obstruction of the intestine in the absence of an anatomical cause and evidence of impaired motility. Given the unusual early-onset CSVD presentation, the first two patients evaluated in the neurology clinic underwent genetic testing for hereditary microangiopathy. Exploratorily, complete ophthalmologic evaluation, including slit lamp biomicroscopy, fundoscopy, orthoptic assessment, fluorescein retinal angiogram, as well as macular, ganglion cell layer and retinal nerve fiber layer optical coherence tomography, was also performed by a neuro-ophthalmologist (E.B.) in the most recently assessed participants (starting June 2025).

### Neuroimaging Assessment

All eligible participants were considered for brain MRI and, except for absolute contraindications, underwent at least one clinically acquired scan using the local standard protocol. A subset of participants underwent serial MRIs (up to four) based on clinical indications and at the discretion of the evaluating neurologist. The most recent MRI, defined as the scan performed closest to the last clinical visit and acquired using the most up-to-date protocol, was independently reviewed by a local neuroradiologist (C.B.) trained in CSVD assessment, in accordance with the Standards for Reporting Vascular Changes on Neuroimaging, version 2 (STRIVE-2) guidelines^10^. Minimal MRI protocol requirements were 1.5 to 3.0T magnetic field strength and acquisition of T1-weighted, T2-weighted, fluid-attenuated inversion recovery (FLAIR), diffusion-weighted (DWI) and T2* or other susceptibility-weighted (either GRE, SWI or SWAN) sequences.

WMH are defined as hyperintense signal on T2-weighted images, including FLAIR, without cavitation. WMH were rated separately in the periventricular and deep white matter of both hemispheres according to the Fazekas scale (regional score range 0-3, total score range 0-6)^11^. Perivascular spaces (PVS) are CSF-isointense, round or linear structures usually ≤2 mm in diameter along penetrating vessels, without FLAIR hyperintensity. Markedly enlarged PVS (>3 mm) were distinguished from lacunes by morphology, surrounding tissue signal, and co-located PVS, in accordance with STRIVE-2. PVS were graded separately in basal ganglia and centrum semiovale on T2-weighted images according to Potter scoring system (regional score range 0-4, with grade 1 for 1-10, grade 2 for 11-20, grade 3 for 21-40 and grade 4 for >40 PVS)^12^. Cortical superficial siderosis (cSS) are thin areas of hypointensity on T2*-weighted and other susceptibility-sensitive sequences located over the superficial cortex. Right and left hemispheres were scored separately for cSS (0 = none, 1 = 1 sulcus or up to 3 immediately adjacent sulci with cSS, and 2 = 2 or more nonadjacent sulci or more than 3 adjacent sulci with cSS) and a total cSS multifocality score (range 0-4) was derived by adding the right and left hemisphere scores^13^. Foci of cSS contiguous or potentially anatomically connected with any lobar intracerebral hemorrhage (ICH) were not included. We provided location of cSS, classified as either supratentorial, infratentorial, or both. Cerebral microbleeds (CMB) are defined as small (≤10 mm), rounded hypointense lesions consistent with hemosiderin deposition identified on susceptibility-sensitive sequences (GRE, SWI or SWAN). We used the Microbleed Anatomical Rating Scale for the assessment of definite microbleeds in the right and left infratentorial, deep and lobar subregions^14^. Lobar regions included the cortex and subcortical white matter (including U fibers), deep regions comprised the basal ganglia, thalamus, internal and external capsules, corpus callosum, and deep/periventricular white matter, and infratentorial regions comprised the brainstem and cerebellum. A total count was provided for definite CMB by adding the right and left hemisphere scores. We also provided a CMB semiquantitative severity score: 0 CMB (grade 0 or absent), 1 or 2 CMB (grade 1 or mild), 3 to 10 CMB (grade 2 or moderate), and >10 (grade 3 or severe)^15–17^. Visual counts and locations were provided for recent small subcortical infarcts (RSSI; DWI-positive lesions of ≤20 mm in diameter within a perforator artery territory), lacunes of presumed vascular origin (round or ovoid, subcortical, CSF-isointense fluid-filled cavity ≤15 mm in diameter), and cortical microinfarcts (CMI; small lesions ≤4 mm confined to the cortex that appear hypointense on T1-weighted, hyperintense on T2/FLAIR, and isointense on T2*-weighted images)^10^. A more global measure of CSVD, the Small Vessel Disease (SVD) score, was derived from imaging results using a previously validated, simple, and pragmatic scale that accounts for the combined presence of lacunes, WMH, CMB, and/or PVS^18^.

Presence and location of other relevant vascular findings included recent and chronic embolic infarcts, acute subarachnoid hemorrhages, and acute, subacute and chronic ICH. Because involvement of anterior temporal lobes and external capsules is characteristic of CADASIL, the prototypical monogenic form of CSVD^19,20^, these regions were also visually inspected for T2/FLAIR hyperintensities. Those for who vascular neuroimaging was also available (either time-of-flight or contrast-enhanced MR-angiogram, or CT angiogram) were additionally assessed for presence and location of intracranial aneurysms and stenoses. Finally, brain atrophy was assessed using the Global Cortical Atrophy (GCA) scale (score range 0-3)^21^, with specification of any lobar predominance when applicable, together with an overall semi-quantitative assessment of subcortical and cerebellar atrophy (regional score range 0-3), based on the neuroradiologist’s global impression.

### Descriptive and Statistical Analyses

Continuous and semi-quantitative variables are summarized as median and range, while categorical variables are summarized as counts and percentages. Semi-quantitative MRI ratings, including WMH, CMB, lacunes, PVS, chronic cerebrovascular lesions and atrophy scores, were analyzed descriptively. Associations between age at MRI and CSVD markers, including CMB severity and summary SVD scores, were assessed using both Spearman’s rank correlation (ρ) and Kendall’s tau (τ) coefficients with 95% bootstrap confidence intervals to account for the small sample size. Further analyses exploring the association between CMB burden and aging were conducted by comparing the mean age of patients with discrete versus countless CMB. Group differences were assessed using Student’s t-test, with normality confirmed by the Shapiro–Wilk test. Two-tailed p-values <0.05 were considered statistically significant. Statistical analyses were conducted in RStudio (http://www.rstudio.com/; R 4.5.2) and GraphPad Prism 10.6.1.

### Standard Protocol Approvals, Registrations, and Patient Consents

The study was approved by the ethics committee of the CHU Sainte-Justine and the CHU de Québec (MP-21-2006-107, F2-29921). Written informed consent for participation in CAID-related research projects was obtained from all patients or their legal representatives.

### Data Availability Statement

Anonymized clinical and derived neuroimaging data are available from the corresponding author upon reasonable request, subject to privacy and ethical restrictions.

### Reporting Guidelines

This manuscript was prepared following STROBE guidelines (**Appendix A**).

## RESULTS

### Participant Clinical Characteristics

The study cohort comprised 16 French-Canadian adult patients with genetically confirmed CAID syndrome, including two pairs of first-degree relatives. Fifteen were homozygous for the *SGO1* mutation and one was a compound heterozygote. The median age at last neurological assessment was 34 years (range, 19–60), with a predominance of women (10/16, 62.5%). A first-degree family history of CAID was reported in seven patients (43.8%). The median age at first clinical manifestation was 13 years (range, 3–38). CIPO was the initial presentation in 13 patients (81.3%), whereas SSS was the first manifestation in three patients (18.8%); all exhibited both CIPO and SSS at genetic confirmation. Fourteen patients (87.5%) had a history of parenteral nutrition, and 12 (75%) were receiving total parenteral nutrition at last follow-up. Eleven patients (68.8%) had undergone at least one abdominal surgery, but none had required intestinal transplantation. Cardiac manifestations included SSS (75%), atrial fibrillation (AF; 50%), atrial flutter (37.5%), atrioventricular block (12.5%), isolated chronotropic incompetence (6.3%), dilated cardiomyopathy (6.3%), bicuspid aortic valve (6.3%), atrial septal defect (12.5%), and ventricular septal defect (6.3%). Thirteen patients (81.3%) had a permanent pacemaker, and nine (56.3%) underwent cardiac interventions, including left atrial appendage (LAA) closure for AF in five (31.3%). The median CHADS₂ score was 0 (range, 0-3). Two patients (12.5%) died during follow-up in their twenties and forties due to nosocomial pneumonia and pulmonary embolism, respectively. No brain autopsy was performed. Detailed demographic and clinical characteristics are summarized in **Table 1**.

**Table 1.**
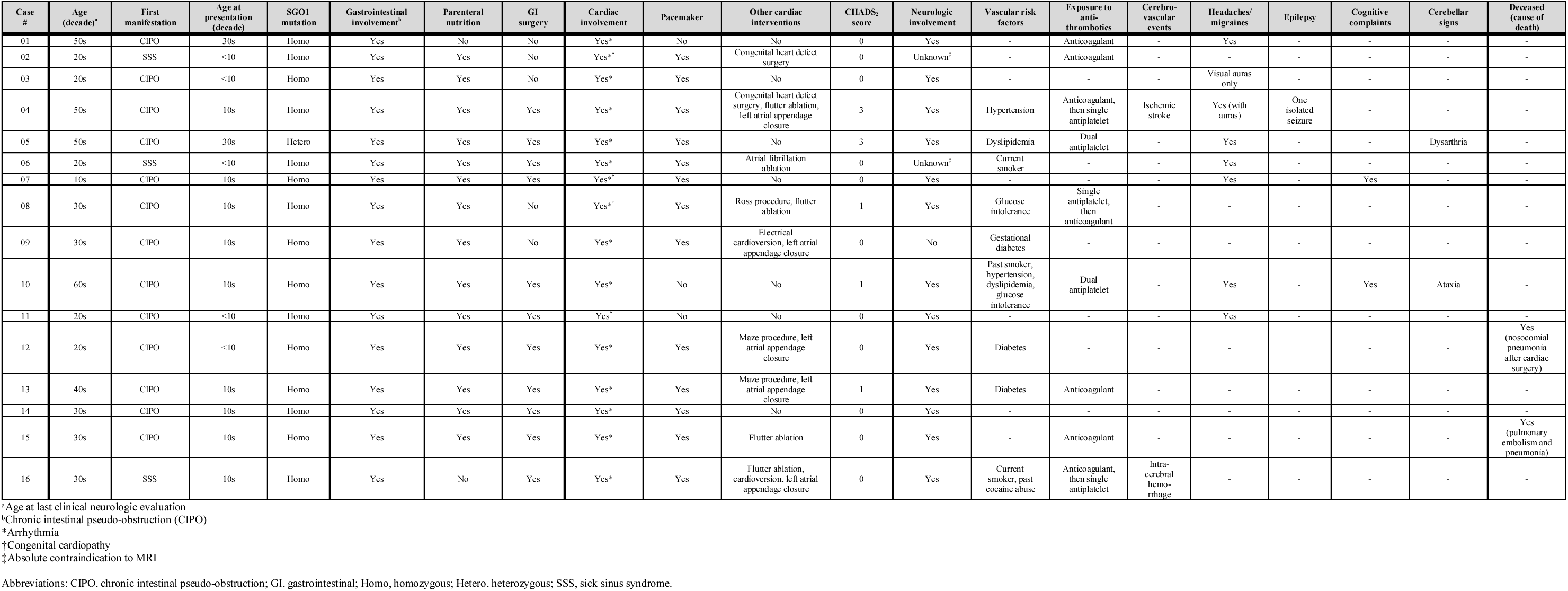
Clinical characteristics of genetically-confirmed cases with chronic atrial and intestinal dysrhythmia (CAID) syndrome.

### Neurological Manifestations

Clinical neurological manifestations potentially related to CSVD were heterogeneous across the cohort. One patient had developmental learning disorders, including dyslexia and dysorthographia, without other neurodevelopmental abnormalities or intellectual disability. A history of migraine was reported in eight patients (44.4%), including migraine with aura in two (11.1%), and isolated recurrent visual auras without headache in one additional case (5.6%). One patient experienced a single unprovoked seizure without subsequent epilepsy. Cognitive complaints were reported by two patients (11.1%), however, no formal cognitive assessment was performed. Cerebellar signs were observed on neurological examination in three patients (16.7%), including gait or limb ataxia in two and dysarthria in one.

In our cohort, two (12.5%) patients experienced a cerebrovascular event. One patient had two simultaneous ischemic strokes, presumed cardioembolic, involving the right occipital lobe and left hippocampus in their early fifties, while another suffered an ICH in their early twenties, attributed to high INR; due to the age of the event, the precise location could not be ascertained from the medical records. Additionally, another patient was evaluated in their forties for transient focal neurological episodes characterized by sensory symptoms, with MRI showing multiple microbleeds but no acute lesions. Importantly, two cases from different families had a brother affected by genetically confirmed CAID syndrome who died from ICH (one cerebellar, one unspecified). Finally, among our CAID participants, two (11.1%) had a history of early-onset sensorineural hearing loss; one (5.6%) had cerebral aneurysms involving the left internal carotid artery and the anterior communicating artery but with a strong family history of cerebral aneurysms in two first- and second-degree relatives; and one (5.6%) had a severe idiopathic axonal sensory neuropathy, all of uncertain significance and association with CAID syndrome.

Three patients from our cohort underwent a comprehensive neuro-ophthalmologic evaluation; the findings are presented in **Appendix B**.

### Brain MRI Findings

Of the 16 participants, 14 underwent brain MRI at the CHU de Québec. The two participants who did not undergo MRI were excluded because of permanent pacemaker incompatibility. Imaging was performed on a 1.5 T system in nine participants (64.3%) and on a 3.0 T system in five (35.7%). All imaged participants (100%) underwent susceptibility-weighted imaging using SWI or SWAN sequences, as well as T1-, T2-, FLAIR, and DWI sequences. Time-of-flight MR angiography was performed in seven participants (50%), and gadolinium contrast–enhanced T1 imaging in three (21.4%). Additionally, three participants (21.4%) underwent CT angiography in the emergency setting.

#### Cerebral Small Vessel Disease

Brain MRI demonstrated an unusually high burden of CSVD in our relatively young sample of adult patients with CAID syndrome (median age 34.5 years, IQR 24–50 years). WMH were present in 92.9% of cases, with median Fazekas scores of 1 (range, 0–3) for both periventricular and deep white matter involvement, and 2 (range, 0–6) for global white matter involvement, including 21.4% with moderate to severe WMH. Anterior temporal lobe and external capsule involvement was found in 14.3% and 42.9% of patients, respectively. Lacunes were observed in 50% of patients, predominantly supratentorial (85.7%), while RSSI and CMI were absent. Moderate-to-severe PVS were identified in 28.6% of cases, with more frequent involvement of basal ganglia compared to centrum semiovale. Notably, although cSS was absent, CMB were highly prevalent, occurring in 85.7% of patients, with involvement of deep, lobar, and infratentorial regions, preferentially affecting the cerebellum (91.7%); five (35.7%) patients exhibited countless CMB (i.e. not quantifiable by visual inspection), and the median CMB severity score was 3 (range, 0– 3). The median summary SVD score was 1.5 (range, 0–4), with only two patients showing no radiological CSVD. Detailed MRI findings can be found in **Table 2** and **Figure 1**. Representative brain MRI images from two selected cases are shown in **Figure 2**, illustrating CSVD lesions of varying severity.

**Figure 1.**
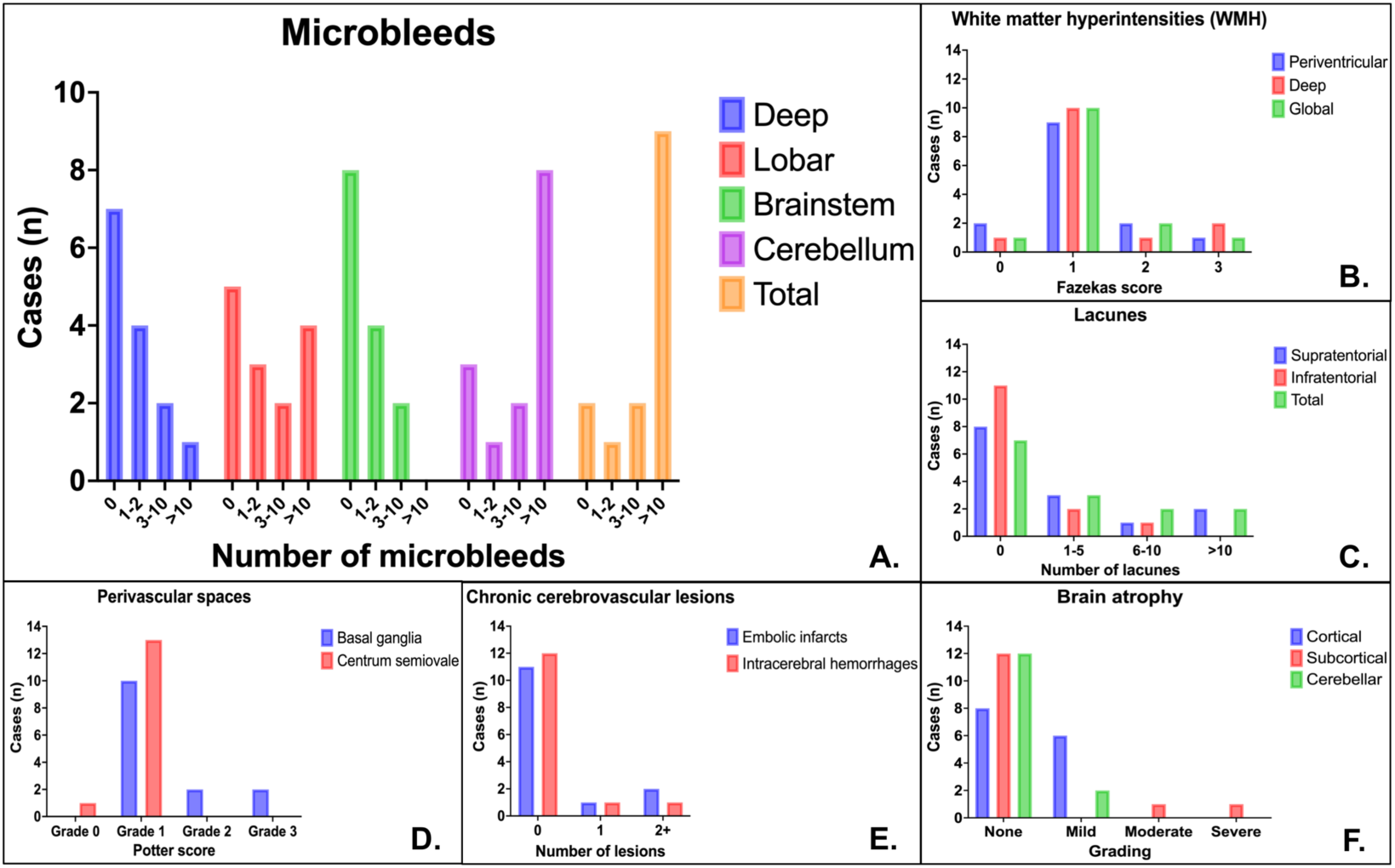
Regional distribution and assessment of MRI markers of CSVD and other cerebrovascular lesions in patients with CAID syndrome. Bar charts show the regional prevalence and severity of A) CMB, B) WMH, C) lacunes of presumed vascular origin, D) PVS, E) chronic cerebrovascular lesions, and F) regional brain atrophy in our cohort of patients with CAID syndrome.

**Figure 2.**
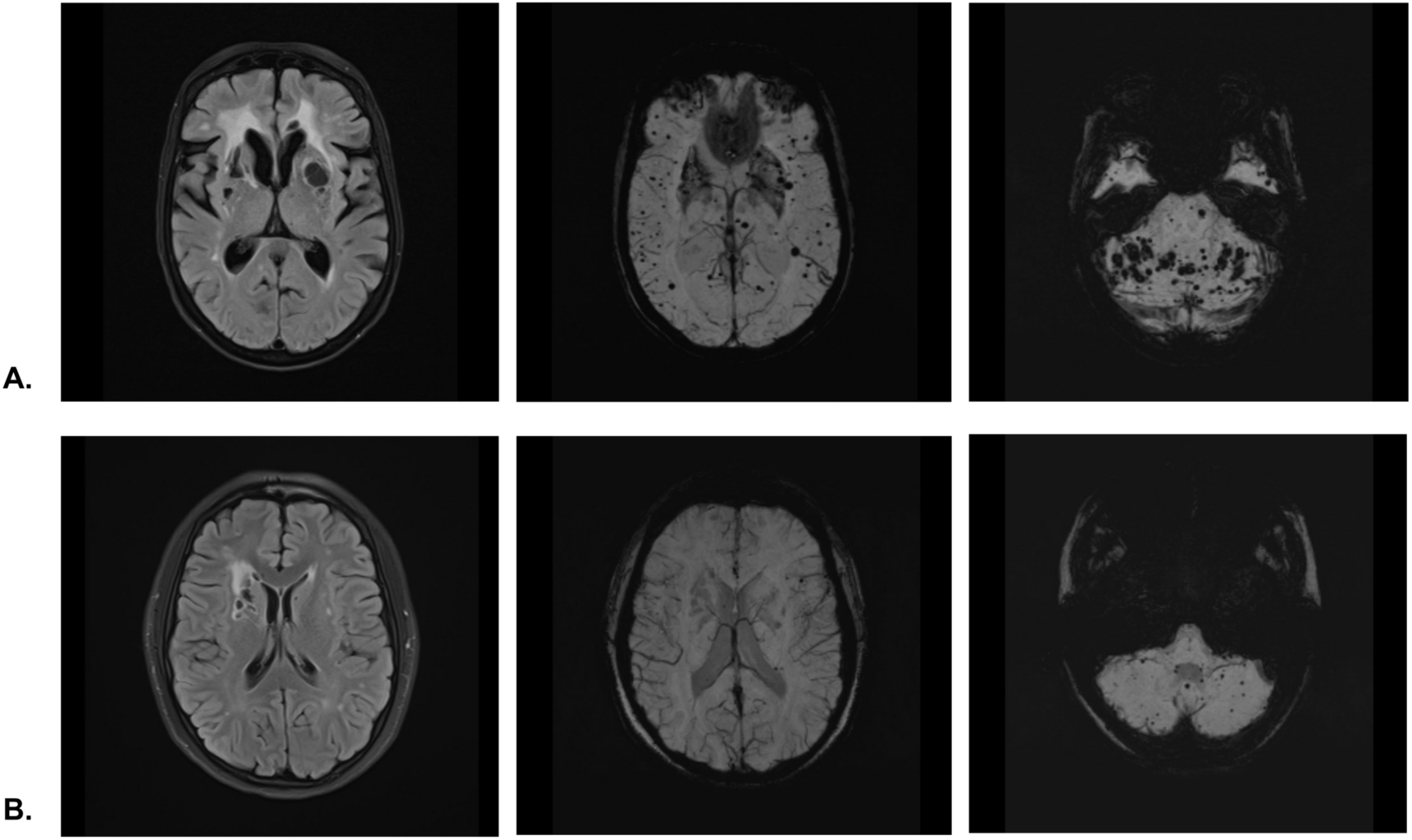
Representative MRI findings of CSVD in CAID syndrome. FLAIR (left) and SWI (middle and right) images from two patients with CAID syndrome in their mid-fifties (A) and early twenties (B). FLAIR images demonstrate diffuse and confluent WMH and multiple deep lacunes of presumed vascular origin. SWI images reveal CMB of varying burden and mixed distribution, predominantly involving the posterior fossa, and particularly the cerebellum.

**Table 2.** Brain MRI findings in cases with chronic atrial and intestinal dysrhythmia (CAID) syndrome.

| Brain MRI Parameters | Prevalence (%) or Median (Range) |
| --- | --- |
| **Magnetic Field Strength** |  |
| 1.5 T | 9/14 (64.3%) |
| 3.0 T | 5/14 (35.7%) |
| **Available Sequences** |  |
| T1 | 14/14 (100%) |
| T2 | 14/14 (100%) |
| FLAIR | 14/14 (100%) |
| DWI | 14/14 (100%) |
| T2*/GRE | 0/14 (0%) |
| SWI/SWAN | 14/14 (100%) |
| Gadolinium-enhanced T1 | 3/14 (21.4%) |
| TOF MRA | 7/14 (50%) |
| Gadolinium-enhanced MRA | 0/14 (0%) |
| **White Matter Hyperintensities (WMH; Fazekas Score)** |  |
| Prevalence (any) | 13/14 (92.9%) |
| Prevalence (grade 2 to 3) | 3/14 (21.4%) |
| Periventricular | 1 (0-3) |
| Deep | 1 (0-3) |
| Global | 1 (0-3) |
| Prevalence of anterior temporal lobe involvement | 2/14 (14.3%) |
| Prevalence of external capsule involvement | 6/14 (42.9%) |
| **Recent Small Subcortical Infarcts** |  |
| Prevalence | 0/14 (0%) |
| Count | 0 (0) |
| Location | NA |
| **Lacunes of Presumed Vascular Origin** |  |
| Prevalence (at least one lacune) | 7/14 (50%) |
| Count | 0.5 (0-20) |
| Location |  |
| Supratentorial | 6/7 (85.7%) |
| Infratentorial | 3/7 (42.9%) |
| **Cortical Microinfarcts** |  |
| Prevalence | 0/14 (0%) |
| Count | 0 (0) |
| Location | NA |
| **Perivascular Spaces (PVS; Potter Scale)** |  |
| Prevalence (moderate-severe) | 4/14 (28.6%) |
| Basal Ganglia | 1 (1-3) |
| Centrum Semiovale | 1 (0-1) |
| **Cortical Superficial Siderosis (cSS; Multifocality Score)** |  |
| Prevalence | 0/14 (0%) |
| Right Hemisphere | 0 (0) |
| Left Hemisphere | 0 (0) |
| Global | 0 (0) |
| Location | NA |
| **Cerebral Microbleeds (CMB; Microbleed Anatomical Rating Scale)** |  |
| Prevalence (at least one CMB) | 12/14 (85.7%) |
| Supratentorial Deep | 7/12 (58.3%) |
| Supratentorial Lobar | 9/12 (75%) |
| Infratentorial Brainstem | 6/12 (50%) |
| Infratentorial Cerebellum | 11/12 (91.7%) |
| Semiquantitative Severity Score (0-3) | 3 (0-3) |
| Cases Described with Countless Microbleeds | 5/14 (35.7%) |
| **Small Vessel Disease Score (SVD; Wardlaw et al.<sup>18</sup>) (0-4)** | 1.5 (0-4) |
| **Other Vascular Findings** |  |
| Recent Embolic Infarcts | 0/14 (0%) |
| Chronic Embolic Infarcts | 3/14 (21.4%) |
| Acute/Subacute Subarachnoid Hemorrhages | 0/14 (0%) |
| Acute/Subacute Intracerebral Hemorrhages | 0/14 (0%) |
| Chronic Intracerebral Hemorrhages | 2/14 (14.3%) |
| Aneurysms | 1/7 (14.3%) |
| Dissections | 0/7 (0%) |
| Stenoses | 0/7 (0%) |
| Gadolinium Enhancement | 1/3 (33.3%) |
| **Brain Atrophy** |  |
| Cortical Atrophy (Global Cortical Atrophy Scale) | 0 (0-1) |
| Lobar Predominance – Temporal | 1/6 (16.7%) |
| Lobar Predominance – Parietal | 3/6 (50%) |
| Lobar Predominance – Temporo-parietal | 2/6 (33.3%) |
| Subcortical Atrophy | 0 (0-3) |
| Cerebellar Atrophy | 0 (0-1) |

Correlations between age at brain MRI and CSVD markers showed weak to moderate positive associations that did not reach statistical significance (**Figure 3**). Specifically, the correlation with CMB severity score was ρ = 0.29 (95% bootstrap CI: −0.23 to 0.71, n = 14), and with summary SVD score was ρ = 0.43 (95% bootstrap CI: −0.27 to 0.83, n = 14). Corresponding Kendall τ coefficients were 0.22 (p = 0.33) and 0.29 (p = 0.17), respectively. While these findings suggest no significant association between age and CSVD burden in this cohort, the mean age was significantly higher in patients with countless CMB than those with discrete CMB (46.2 ± 15.7 vs. 31.2 ± 9.4 years; t₁₂ = -2.26, p = 0.043, 95% CI for the mean difference: -29.4 to -0.53; **Appendix C**). These results suggest that patients with countless CMB tend to be older than those with a quantifiable CMB burden, suggesting a potential association with aging. However, these results must be interpreted with caution due to the limited sample size.

**Figure 3.**
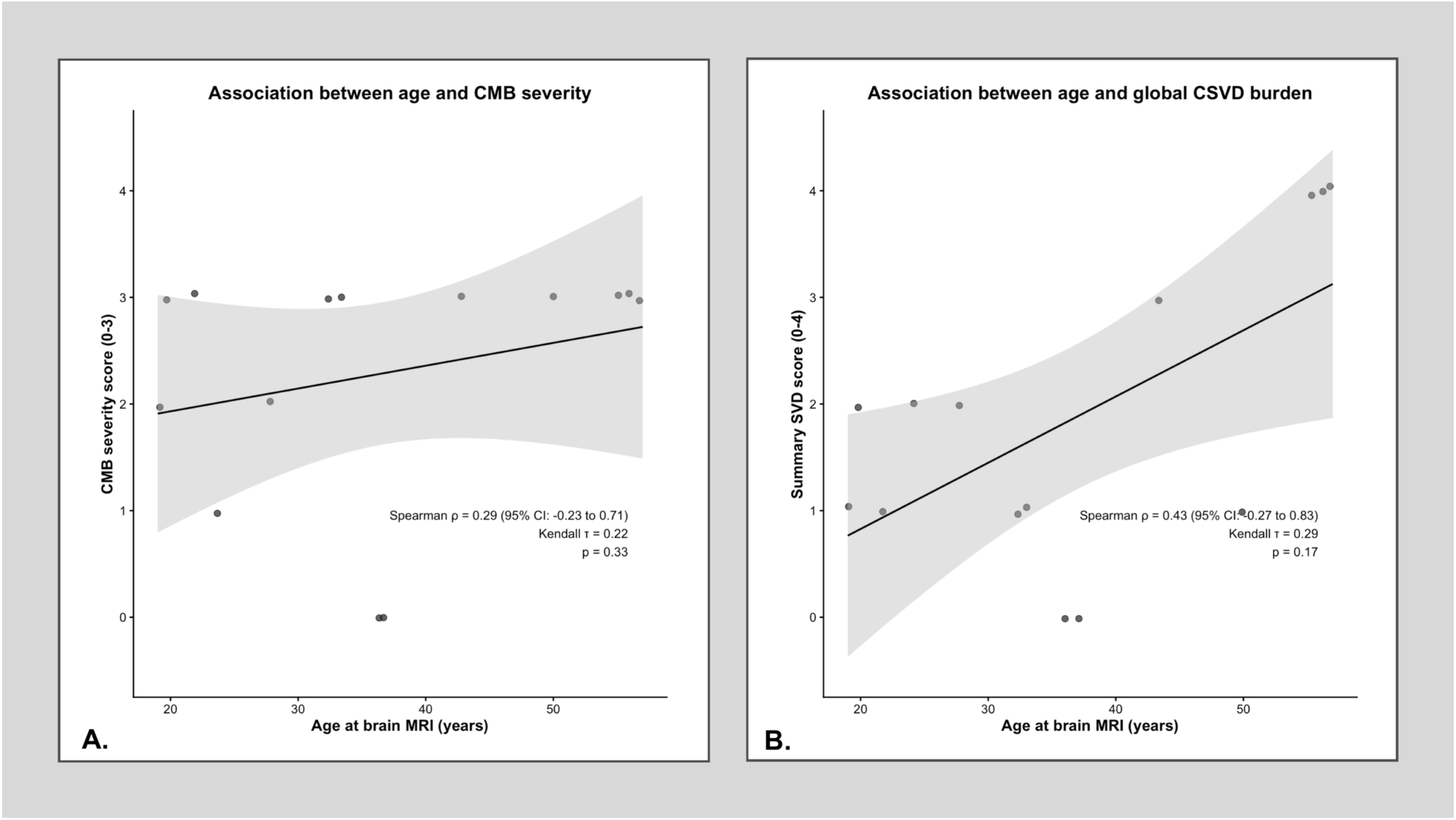
Relationship between age and CSVD lesions in CAID syndrome. Scatterplots showing unadjusted correlations between age at brain MRI and (A) global CMB severity and (B) CSVD burden scores. Spearman’s rank correlation (ρ, 95% CI) and Kendall’s tau (τ) coefficients with corresponding p-values are indicated.

#### Macrovascular Lesions

Additional vascular findings included chronic embolic infarcts in three (21.4%) participants and chronic ICH in two (14.3%) participants. No participants showed evidence of recent embolic infarcts or acute/subacute subarachnoid hemorrhage or ICH. Among participants who underwent vascular imaging, no cerebral aneurysms were identified, with the exception of one participant with a previously treated aneurysm who had undergone surgical occlusion of the left internal carotid artery. No intra- or extracranial arterial stenosis or dissection was observed.

#### Other Findings

Aside from vascular-specific lesions, one participant demonstrated unusual multifocal parenchymal gadolinium enhancement, with multiple foci involving the left lenticular nucleus and both cerebellar hemispheres (**Figure 4**). Brain atrophy was generally mild, with a median Global Cortical Atrophy Scale score of 0 (range, 0–1). Mild cortical atrophy was present in six participants (42.6%), most commonly with parietal predominance (50%), followed by temporoparietal (33.3%) and temporal (16.7%) involvement. Subcortical atrophy was observed in two patients (14.3%), graded as moderate in one case and severe in the other, while cerebellar atrophy was minimal overall; only these two patients also exhibited concomitant mild cerebellar atrophy. Median scores for subcortical and cerebellar atrophy were 0 (ranges 0–3 and 0–1, respectively). These neuroimaging findings are detailed in **Table 2**.

**Figure 4.**
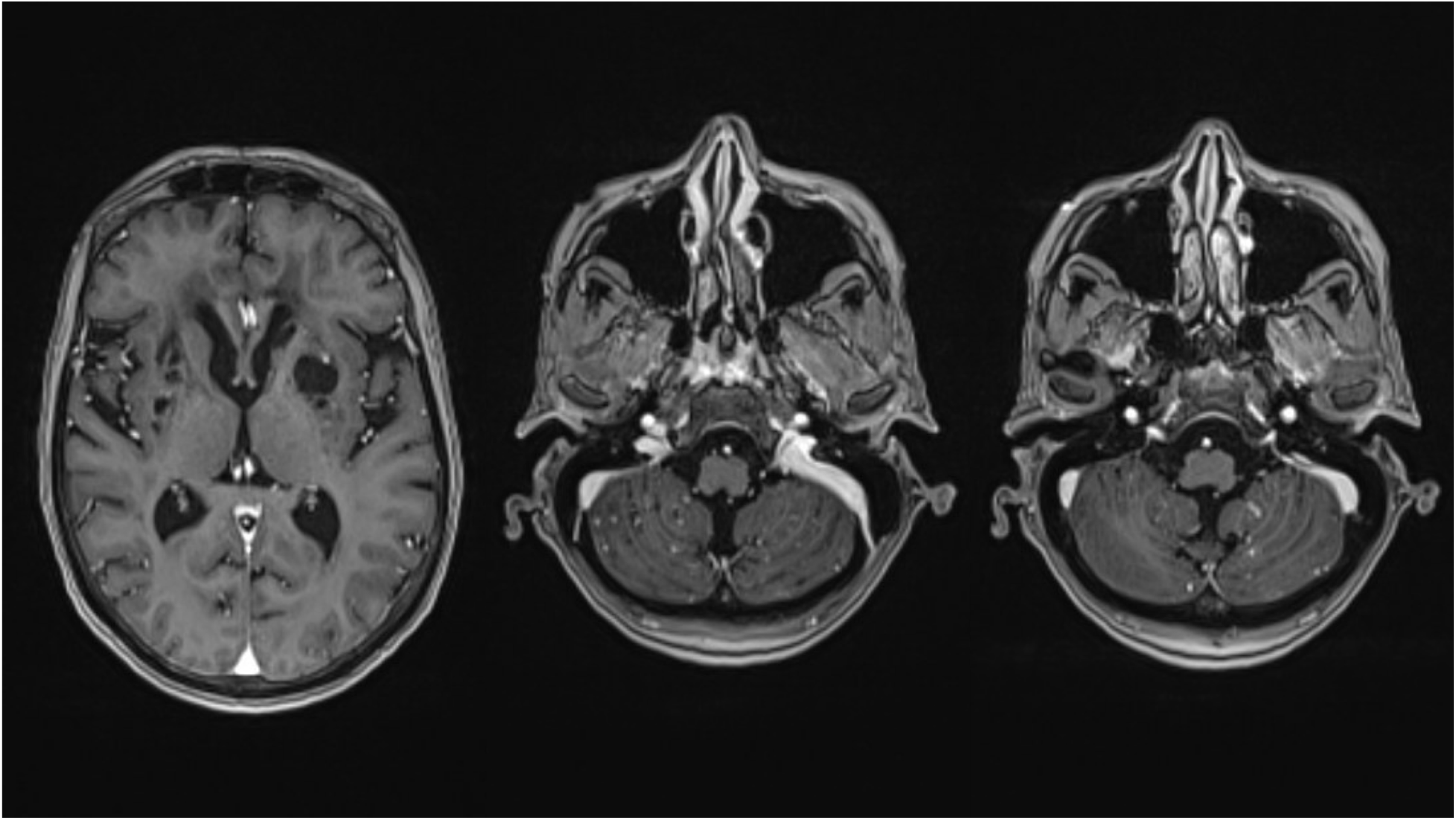
Unusual multifocal parenchymal gadolinium enhancement in CAID syndrome. Post-contrast T1-weighted MRI images from a patient in their mid-fifties (same patient as in Figure 2A) show multiple enhancing foci in the left lenticular nucleus and both cerebellar hemispheres, illustrating atypical multifocal parenchymal involvement.

### Management of Cardioembolic Risk

Among the eight CAID patients with AF, cardioembolic risk management was individualized according to CHADS₂ score^22^, hemorrhagic risk, and neuroimaging findings. One patient with a higher-risk CHADS₂ score of 3 was cautiously treated with reduced-dose apixaban before undergoing LAA closure, followed by aspirin. Two patients with a score of 1 underwent LAA closure; one subsequently received long-term dual antiplatelet therapy for peripheral artery disease, while the other, previously on vitamin K antagonists and low-molecular-weight heparin, remained on anticoagulation due to recurrent deep vein thrombosis. The remaining patients (n=5) had CHADS₂ scores of 0 and were mostly managed conservatively, either without antithrombotic therapy (n=2) or with anticoagulation prescribed for non-cardioembolic indications such as venous thromboembolism (n=1). In two cases, LAA closure was nevertheless performed, either in the context of another planned cardiac procedure (n=1) or because of a complex cardiac condition with severe atrial disease conferring a high cardioembolic risk (n=1). When not contraindicated, MRI findings, particularly CMB burden, influenced management decisions, with high CMB burden favoring avoidance of long-term anticoagulation. Overall, LAA closure was frequently employed as an alternative strategy to mitigate cardioembolic risk while limiting hemorrhagic complications in this high-risk population. Details of local cardiac surgical strategies in patients with CAID syndrome are provided in **Appendix D**.

## DISCUSSION

### Summary of Results

In this genetically confirmed cohort of 16 adult patients with CAID syndrome due to an autosomal recessive *SGO1* mutation, typically characterized by a unique association of gastrointestinal and cardiac manifestations, we identified a striking burden of CSVD despite the relatively young age of the participants. Brain MRI revealed a high prevalence of WMH, lacunes, and especially CMB, frequently involving multiple territories, including deep, lobar, and infratentorial regions, with a unique marked cerebellar predominance. Cerebrovascular events were relatively uncommon but included both ischemic stroke and ICH, while transient focal neurological symptoms and family history of ICH in affected relatives further support an increased cerebrovascular risk in these patients. Additional neurological manifestations included migraine and mild cerebellar signs. In this rare population, characterized by both a high incidence of congenital cardiopathies and arrhythmias and an unusually high CMB burden, AF management required highly individualized strategies. The frequent use of LAA closure reflected the complex balance between stroke prevention and hemorrhagic risk.

### Evidence in Context

#### A Monogenic Cause of Autosomal Recessive CSVD

Our findings suggest that CAID syndrome is a likely monogenic cause of CSVD, expanding the spectrum of genetically mediated CSVD beyond well-recognized conditions such as CADASIL, COL4A1/2, or HTRA1-related disorders (**Table 3**)^19,20^. In patients presenting with young-onset CSVD alongside systemic features, particularly gastrointestinal dysmotility and congenital cardiac abnormalities or arrhythmia, CAID syndrome should be considered in the differential diagnosis. This aligns with previous reports by *Nehme et al.*^4^, describing two cases of CAID syndrome presenting with neurological symptoms (one ICH and one ischemic stroke) alongside normal neuro-ophthalmologic examinations, normal CSF analyses, and negative conventional hereditary microangiopathy panels, and by *Schuermans et al*.^5^, who reported a young affected adult with recurrent ICH and extensive WMH with diffuse cerebellar CMB. Whether CSVD represents an early or late manifestation of the disease remains uncertain, as brain MRI is rarely performed in asymptomatic children, and no prior studies have specifically addressed this question. Nevertheless, the substantial burden of CMB observed in our relatively young cohort, including patients in their early twenties, supports the plausibility of subclinical cerebrovascular involvement beginning early in life, independent of gastrointestinal or cardiac manifestations. This novel association of a *SGO1*-related systemic disorder with concomitant neurological involvement is perhaps unsurprising, as *SGO1* is robustly expressed in the neural tube during mammalian development in mouse models, suggesting a direct role in cerebral development^23^. Reflecting the involvement of pacemaker and autonomic cells in the gastrointestinal and cardiac systems, a compelling hypothesis is that cerebral microvascular injury stems from dysfunction of vascular smooth muscle cells, which are critical for vessel wall integrity and cerebral autoregulation. Another plausible mechanism involves intrinsic endothelial defects, resulting in endothelial dysfunction and compromised vascular integrity, potentially compounded by *SGO1* mutation-associated premature vascular senescence and aging, which may also underlie the perivascular parenchymal enhancement observed in one patient. Additional mechanisms, including amyloid-related vascular changes^24^ and dysregulated TGF-β signaling^3,4^—which has been implicated in multiple forms of CSVD, including CADASIL^25^—may underlie the pronounced microvascular vulnerability observed in CAID patients and also warrant investigation. With aging, posterior circulation arteries have thinner walls and less elastin than anterior vessels^26^, making them particularly vulnerable to injury; in CAID, this may be exacerbated by premature senescence of cerebellar arterioles and TGF-β–induced vessel wall fibrosis, which together could potentially explain the observed predilection for CSVD lesions in the posterior fossa and cerebellum. Histopathological and biomarker studies will be essential to clarify the precise vascular injury pathways in CAID syndrome.

**Table 3.** Well-Established Monogenic Causes of Cerebral Small Vessel Disease.

| Name | Gene | Inheritance | Clinical Manifestations | Distinctive Neuroimaging Features |
| --- | --- | --- | --- | --- |
| CADASIL | NOTCH3 | AD | Migraine with aura, ischemic stroke, cognitive decline, psychiatric symptoms | Confluent WMH with anterior temporal pole and external capsule involvement |
| CARASIL | HTRA1 | AR | Early-onset ischemic stroke, cognitive decline, alopecia, premature cervical or lumbosacral spondylosis | Diffuse WMH, high T2 signal from the pons to the middle cerebellar peduncles (arc sign) |
| HTRA1-related AD CSVD | HTRA1 | AD | Milder CARASIL phenotype with later onset | Diffuse supratentorial and pontine WMH |
| PADMAL | COL4A1 | AD | Recurrent pontine ischemic strokes, dysarthria, gait impairment | Confluent WMH and lacunar infarcts predominantly involving the pons |
| CARASAL | CTSA | AD | Stroke, therapy-resistant hypertension, hearing loss, cognitive decline | CADASIL-mimicking WMH with early pontine involvement |
| LAMB1-related AD leukoencephalopathy | LAMB1 | AD | Episodic (hippocampic) memory impairment, seizures and stroke-like episodes | Diffuse WMH, cortical/cerebellar dysplasia and cysts |
| FOXC1 deletion-related angiopathy | FOXC1 | AD | Stroke, cognitive decline, and ocular abnormalities (particularly anterior segment dysgenesis/Axenfeld-Rieger syndrome) | WMH, vertebrobasilar dolichoectasia, ventriculomegaly, cerebellar hypoplasia |
| COL4A1/COL4A2-related microangiopathy | COL4A1, COL4A2 | AD | ICH, eye disease (retinal arteriolar tortuosity, cataracts, anterior segment defects), kidney disease, Raynaud phenomenon, muscle cramps, supraventricular arrhythmia | Deep CMB, porencephalic cysts |
| Hereditary cerebral amyloid angiopathy | APP, CST3, GSN, TTR, ITM2B | AD | ICH, cognitive decline, seizures (variant-dependent) | Lobar CMB, cortical subarachnoid hemorrhage and superficial siderosis |
| RVCL-S | TREX1 | AD | Stroke, retinal vasculopathy (hemorrhages and cotton wool spots), cognitive decline, microvascular liver and kidney disease, arterial hypertension, Raynaud phenomenon | Multifocal enhancing white matter lesions, punctate diffusion restriction, brain calcifications and pseudotumoral lesions |
| Leukoencephalopathy with cysts and calcifications | SNORD118 | AR | Cognitive decline, seizures, movement disorders | Triad of diffuse WMH, intracranial calcifications, and parenchymal cysts |
| Fabry disease | GLA | X-linked | Hypertrophic cardiomyopathy, nephropathy with proteinuria, acroparesthesia, angiokeratomas, corneal and lenticular opacities | WMH (especially frontal and parietal lobes), pulvinar sign, vertebrobasilar dolichoectasia |

**Abbreviations:** AD, autosomal dominant; AR, autosomal recessive; CADASIL, cerebral autosomal dominant arteriopathy with subcortical infarcts and leukoencephalopathy; CARASIL, cerebral autosomal recessive arteriopathy with subcortical infarcts and leukoencephalopathy; CMB, cerebral microbleeds; CSVD, cerebral small vessel disease; ICH, intracerebral hemorrhage; PADMAL, pontine autosomal dominant microangiopathy and leukoencephalopathy; RVCL-S, retinal vasculopathy with cerebral leukodystrophy and systemic manifestations; WMH, white matter hyperintensities.

#### Clinical Correlation of Neurologic Involvement in CAID Syndrome

Neurologic involvement in CAID syndrome appears to manifest primarily in adulthood and is distinct from other cohesinopathies, as patients generally do not exhibit neurodevelopmental anomalies, intellectual disability, or epilepsy. Clinically, the spectrum includes mild cerebellar signs, transient focal neurological episodes, and an increased risk of cerebrovascular events. Headache and migraine were reported in a subset of patients, suggesting possible overlap with other monogenic CSVD such as CADASIL. Cognitive impairment may also occur, raising the concern for early-onset dementia in CAID syndrome, particularly given the radiologic resemblance of some lesions to those seen in cerebral amyloid angiopathy^27^ and other CSVD subtypes associated with vascular cognitive impairment^28^. While cerebral aneurysms have been reported in the literature and one case was observed in our cohort, their significance remains uncertain. Overall, the neurological phenotype in CAID seems to be predominantly adult-onset, although subclinical or radiologic manifestations in pediatric patients cannot yet be excluded.

#### A Population at High Risk for Both Ischemic and Hemorrhagic Stroke

Patients with CAID syndrome represent a population at uniquely high risk for both ischemic and hemorrhagic cerebrovascular events. Congenital cardiopathies and arrhythmias, including AF and sinus node dysfunction, predispose to cardioembolic strokes, while the striking burden of CSVD and CMB—together with prior and future necessary cardiac surgeries, parenteral nutrition catheter–related thromboses requiring anticoagulation, and occasional aneurysms—elevates the risk of ICH. This dual vulnerability poses significant challenges for clinical management, particularly regarding the use of anticoagulant and other antithrombotic therapies, requiring careful balancing of ischemic versus hemorrhagic risk in a manner reminiscent of strategies employed in cerebral amyloid angiopathy.

#### Proposed Management of CAID Patients from a Neurologist’s Perspective

Given the high prevalence of associated CSVD, a comprehensive neurological evaluation and brain imaging, including susceptibility-sensitive sequences, may be valuable upon genetic confirmation of CAID syndrome. Additional critical time points at which brain MRI should be considered include prior to initiating anticoagulation for AF or thrombosis, or prior to pacemaker implantation, to better understand baseline cerebrovascular burden and inform risk assessment. In selected cases, prophylactic LAA closure might be considered, particularly if cardiac surgery is planned or if AF coexists with a high burden of CMB. Considering the anticipated need for longitudinal neuroimaging, an MRI-conditional pacemaker should be preferred. As a complex multisystemic disease, optimal care requires a multidisciplinary, consensus-driven approach involving neurologists, cardiologists and gastroenterologists collaborating alongside other healthcare professionals to manage the full spectrum of comorbidities. Primary prevention through careful management of vascular risk factors, as well as tailored symptomatic treatment of headaches while accounting for cerebrovascular vulnerability, may further benefit this population. Such a proactive, individualized strategy is essential to balance ischemic and hemorrhagic risk in this vulnerable population.

### Limitations

This study represents the first comprehensive description of neurological involvement in a relatively large cohort affected by this rare multisystemic disorder. Indeed, CAID syndrome is a recently recognized genetic condition, first described in 2014^1^, for which knowledge is still emerging. Our findings provide compelling evidence of an existing CAID-associated CSVD, emphasizing the need for recognition by the medical community and for detailed phenotyping of affected individuals. Nevertheless, several limitations must be acknowledged. The sample size was modest, which inherently limited statistical power and constrained the scope of analyses, an issue unavoidable in the context of such a rare disease. Neuro-ophthalmologic evaluations were not systematically performed, and formal cognitive testing was not conducted, which may have underestimated the full spectrum of neurological involvement. Biofluid biomarkers and postmortem data were not available, precluding biological or neuropathological confirmation. Additionally, the study was retrospective, monocentric, and lacked a control group, which limits direct inference about disease progression and specificity. The retrospective nature of the study also precluded detailed characterization of some clinical features, including migraine aura and severity. Despite these constraints, the findings provide valuable insights into the neurological phenotype of CAID syndrome and lay the groundwork for future prospective, multicenter studies.

## CONCLUSION

CAID syndrome due to biallelic *SGO1* mutations appears to represent a novel monogenic cause of CSVD, characterized by an unexpectedly high CMB burden despite a young age, particularly in the posterior circulation. This marked cerebrovascular vulnerability coexists with congenital cardiac disease and arrhythmias, placing patients at uniquely high risk for both ischemic and hemorrhagic stroke. Our findings underscore the importance of systematic neurological assessment and brain MRI in the clinical management of CAID patients, particularly when considering antithrombotic strategies or cardiac interventions. Further multicenter and longitudinal studies are needed to clarify disease mechanisms, natural history, and optimal preventive strategies in this rare but potentially high-risk population.

## Supplementary Data

Online access to Supplemental Material is available at [x].

## Acknowledgments

C.D.T., A.N., and S.V. designed the study. P.C., G.A., and S.V. recruited CAID syndrome participants. Neurological assessments were conducted by C.D.T., M.C.C., and S.V. C.D.T. and F.B. collected clinical data from medical records. C.D.T. developed the neuroimaging CSVD assessment protocol and data collection tools. C.B. performed MRI reading and rating, and E.B. conducted the neuro-ophthalmological examinations. C.D.T. analyzed and interpreted the data and drafted the manuscript. A.N., M.B., A.D., P.C., G.A. and S.V. provided critical revision of the final manuscript for important intellectual content. All other authors critically revised the manuscript and approved the final version. We gratefully thank the patients with CAID syndrome who generously consented to participate in this study; their invaluable contribution is fundamental to advancing research on this rare genetic disease.

## ABBREVIATIONS

AF: atrial fibrillation
CADASIL: cerebral autosomal dominant arteriopathy with subcortical infarcts and leukoencephalopathy
CAID: Chronic atrial and intestinal dysrhythmia
CIPO: chronic intestinal pseudo-obstruction
CMB: cerebral microbleeds
CMI: cortical microinfarcts
CSF: cerebrospinal fluid
cSS: cortical superficial siderosis
CSVD: cerebral small vessel disease
CT: computed tomography
DWI: diffusion-weighted imaging
FLAIR: fluid-attenuated inversion recovery
GCA: Global Cortical Atrophy
GRE: gradient-recalled echo
ICH: intracerebral hemorrhage
LAA: left atrial appendage
MARS: Microbleed Anatomical Rating Scale
MRI: magnetic resonance imaging
PVS: perivascular spaces
RSSI: recent small subcortical infarcts
SSS: sick sinus syndrome
STRIVE-2: Standards for Reporting Vascular Changes on Neuroimaging, version 2
SVD: Small Vessel Disease
SWAN: susceptibility-weighted angiography
SWI: susceptibility weighted imaging
WMH: white matter hyperintensities.

## REFERENCES

1. Chetaille P, Preuss C, Burkhard S, Cote JM, Houde C, Castilloux J, Piche J, Gosset N, Leclerc S, Wunnemann F, et al. Mutations in SGOL1 cause a novel cohesinopathy affecting heart and gut rhythm. Nat Genet. 2014;46:1245–1249. doi: 10.1038/ng.3113

2. Piche J, Van Vliet PP, Puceat M, Andelfinger G. The expanding phenotypes of cohesinopathies: one ring to rule them all! Cell Cycle. 2019;18:2828–2848. doi: 10.1080/15384101.2019.1658476

3. Piche J, Gosset N, Legault LM, Pacis A, Oneglia A, Caron M, Chetaille P, Barreiro L, Liu D, Qi X, et al. Molecular Signature of CAID Syndrome: Noncanonical Roles of SGO1 in Regulation of TGF-beta Signaling and Epigenomics. Cell Mol Gastroenterol Hepatol. 2019;7:411–431. doi: 10.1016/j.jcmgh.2018.10.011

4. Nehme A, Gioia LC, Jacquin G, Poppe AY, Letourneau-Guillon L, Odier C. Chronic Atrial Intestinal Dysrhythmia Syndrome Is Associated with Cerebral Small Vessel Disease and Predominantly Cerebellar Microbleeds. Can J Neurol Sci. 2020;47:566–568. doi: 10.1017/cjn.2020.57

5. Schuermans N, Hemelsoet D, Terryn W, Steyaert S, Van Coster R, Coucke PJ, Steyaert W, Callewaert B, Bogaert E, Verloo P, et al. Shortcutting the diagnostic odyssey: the multidisciplinary Program for Undiagnosed Rare Diseases in adults (UD-PrOZA). Orphanet J Rare Dis. 2022;17:210. doi: 10.1186/s13023-022-02365-y

6. Yadav A, Garg AK, Veerwal H, Bhatia P, Bhattacharya A, Sharma V. When gut meets the heart: Chronic atrial and intestinal dysrhythmia presenting as chronic intestinal pseudo-obstruction, an uncommon cohesinopathy. Indian J Gastroenterol. 2024;43:1223–1225. doi: 10.1007/s12664-024-01521-5

7. Zengin O, Gore B, Sahiner ES, Ates I. CAID syndrome and a two year clinical outcome. A case report. J Gastrointestin Liver Dis. 2024;33:571–572. doi: 10.15403/jgld-5772

8. Ahuja K, Pathania S, Baron N, Khlevner J, Bialer M, Mait-Kaufman J. Chronic atrial and intestinal dysrhythmia: A rare genetic disorder of intestinal pseudo-obstruction. JPGN Rep. 2025;6:162–165. doi: 10.1002/jpr3.12158

9. Parelho V, Hadjur S, Spivakov M, Leleu M, Sauer S, Gregson HC, Jarmuz A, Canzonetta C, Webster Z, Nesterova T, et al. Cohesins functionally associate with CTCF on mammalian chromosome arms. Cell. 2008;132:422–433. doi: 10.1016/j.cell.2008.01.011

10. Duering M, Biessels GJ, Brodtmann A, Chen C, Cordonnier C, de Leeuw FE, Debette S, Frayne R, Jouvent E, Rost NS, et al. Neuroimaging standards for research into small vessel disease-advances since 2013. Lancet Neurol. 2023;22:602–618. doi: 10.1016/S1474-4422(23)00131-X

11. Fazekas F, Chawluk JB, Alavi A, Hurtig HI, Zimmerman RA. MR signal abnormalities at 1.5 T in Alzheimer’s dementia and normal aging. AJR Am J Roentgenol. 1987;149:351–356. doi: 10.2214/ajr.149.2.351

12. Potter GM, Chappell FM, Morris Z, Wardlaw JM. Cerebral perivascular spaces visible on magnetic resonance imaging: development of a qualitative rating scale and its observer reliability. Cerebrovasc Dis. 2015;39:224–231. doi: 10.1159/000375153

13. Charidimou A, Boulouis G, Roongpiboonsopit D, Auriel E, Pasi M, Haley K, van Etten ES, Martinez-Ramirez S, Ayres A, Vashkevich A, et al. Cortical superficial siderosis multifocality in cerebral amyloid angiopathy: A prospective study. Neurology. 2017;89:2128–2135. doi: 10.1212/WNL.0000000000004665

14. Gregoire SM, Chaudhary UJ, Brown MM, Yousry TA, Kallis C, Jager HR, Werring DJ. The Microbleed Anatomical Rating Scale (MARS): reliability of a tool to map brain microbleeds. Neurology. 2009;73:1759–1766. doi: 10.1212/WNL.0b013e3181c34a7d

15. Kim HS, Lee DH, Ryu CW, Lee JH, Choi CG, Kim SJ, Suh DC. Multiple cerebral microbleeds in hyperacute ischemic stroke: impact on prevalence and severity of early hemorrhagic transformation after thrombolytic treatment. AJR Am J Roentgenol. 2006;186:1443–1449. doi: 10.2214/AJR.04.1933

16. Shoamanesh A, Pearce LA, Bazan C, Catanese L, McClure LA, Sharma M, Marti-Fabregas J, Anderson DC, Kase CS, Hart RG, et al. Microbleeds in the Secondary Prevention of Small Subcortical Strokes Trial: Stroke, mortality, and treatment interactions. Ann Neurol. 2017;82:196–207. doi: 10.1002/ana.24988

17. Shoamanesh A, Kwok CS, Lim PA, Benavente OR. Postthrombolysis intracranial hemorrhage risk of cerebral microbleeds in acute stroke patients: a systematic review and meta-analysis. Int J Stroke. 2013;8:348–356. doi: 10.1111/j.1747-4949.2012.00869.x

18. Staals J, Makin SD, Doubal FN, Dennis MS, Wardlaw JM. Stroke subtype, vascular risk factors, and total MRI brain small-vessel disease burden. Neurology. 2014;83:1228–1234. doi: 10.1212/WNL.0000000000000837

19. Manini A, Pantoni L. Genetic Causes of Cerebral Small Vessel Diseases: A Practical Guide for Neurologists. Neurology. 2023;100:766–783. doi: 10.1212/WNL.0000000000201720

20. Guey S, Chabriat H. Monogenic causes of cerebral small vessel disease and stroke. Handb Clin Neurol. 2024;204:273–287. doi: 10.1016/B978-0-323-99209-1.00018-1

21. Harper L, Barkhof F, Fox NC, Schott JM. Using visual rating to diagnose dementia: a critical evaluation of MRI atrophy scales. J Neurol Neurosurg Psychiatry. 2015;86:1225–1233. doi: 10.1136/jnnp-2014-310090

22. Gage BF, Waterman AD, Shannon W, Boechler M, Rich MW, Radford MJ. Validation of clinical classification schemes for predicting stroke: results from the National Registry of Atrial Fibrillation. JAMA. 2001;285:2864–2870. doi: 10.1001/jama.285.22.2864

23. Song AT, Galli A, Leclerc S, Nattel S, Mandato C, Andelfinger G. Dataset of Sgo1 expression in cardiac, gastrointestinal, hepatic and neuronal tissue in mouse. Data Brief. 2017;13:731–737. doi: 10.1016/j.dib.2017.06.046

24. Rao CV, Farooqui M, Asch AS, Yamada HY. Critical role of mitosis in spontaneous late-onset Alzheimer’s disease; from a Shugoshin 1 cohesinopathy mouse model. Cell Cycle. 2018;17:2321–2334. doi: 10.1080/15384101.2018.1515554

25. Heidari P, Taghizadeh M, Vakili O. Signaling pathways and molecular mechanisms involved in the onset and progression of cerebral autosomal dominant arteriopathy with subcortical infarcts and leukoencephalopathy (CADASIL); a focus on Notch3 signaling. J Headache Pain. 2025;26:96. doi: 10.1186/s10194-025-02025-z

26. Roth W, Morgello S, Goldman J, Mohr JP, Elkind MS, Marshall RS, Gutierrez J. Histopathological Differences Between the Anterior and Posterior Brain Arteries as a Function of Aging. Stroke. 2017;48:638–644. doi: 10.1161/STROKEAHA.116.015630

27. Charidimou A, Boulouis G, Frosch MP, Baron JC, Pasi M, Albucher JF, Banerjee G, Barbato C, Bonneville F, Brandner S, et al. The Boston criteria version 2.0 for cerebral amyloid angiopathy: a multicentre, retrospective, MRI-neuropathology diagnostic accuracy study. Lancet Neurol. 2022;21:714–725. doi: 10.1016/S1474-4422(22)00208-3

28. VasCog WSOCC, Sachdev PS, Bentvelzen AC, Kochan NA, Jiang J, Hosoki S, Koncz R, Chander RJ, Saks D, Aben HP, et al. Revised Diagnostic Criteria for Vascular Cognitive Impairment and Dementia-The VasCog-2-WSO Criteria. JAMA Neurol. 2025;82:1103–1112. doi: 10.1001/jamaneurol.2025.3242

