## Supplemental Material for "Chronic Atrial and Intestinal Dysrhythmia Syndrome: A Distinct Monogenic Cause of Cerebral Small Vessel Disease"

### Appendix A – STROBE Statement: Checklist of items that should be included in reports of *cross-sectional studies*

| | Item Description | Location (or reason for not reporting) |
|---|---|---|
| **Title and abstract** | | |
| 1a. Indicate the study's design | Indicate the study's design with a commonly used term in the title or the abstract. | Abstract; Page 4 |
| 1b. Abstract | Provide in the abstract an informative and balanced summary of what was done and what was found. | Abstract; Page 4 |
| **Introduction** | | |
| 2. Background / rationale | Explain the scientific background and rationale for the investigation being reported. | Introduction; Pages 6-7 |
| 3. Objectives | State specific objectives, including any prespecified hypotheses. | Introduction; Page 7 |
| **Methods** | | |
| 4. Study design | Present key elements of study design early in the paper. | Methods; Page 7 |
| 5. Setting | Describe the setting, locations, and relevant dates, including periods of recruitment, exposure, follow-up, and data collection. | Methods; Page 7 |
| 6a. Eligibility criteria | **Cohort study:** Give the eligibility criteria, and the sources and methods of selection of participants. Describe methods of follow-up. **Case-control study:** Give the eligibility criteria, and the sources and methods of case ascertainment and control selection. Give the rationale for the choice of cases and controls. **Cross-sectional study:** Give the eligibility criteria, and the sources and methods of selection of participants. | Methods; Page 7 |
| 6b. Matching criteria | **Cohort study:** For matched studies, give matching criteria and number of exposed and unexposed. **Case-control study:** For matched studies, give matching criteria and the number of controls per case. | N/A |
| 7. Variables | Clearly define all outcomes, exposures, predictors, potential confounders, and effect modifiers. Give diagnostic criteria, if applicable. | Methods; Pages 7-10 |
| 8. Data sources / measurement | For each variable of interest give sources of data and details of methods of assessment (measurement). Describe comparability of assessment methods if there is more than one group. | Methods; Page 7-10 |
| 9. Bias | Describe any efforts to address potential sources of bias. | Methods; Pages 7-10 |
| 10. Study size | Explain how the study size was arrived at. | Methods; Page 7 |

| | | |
|---|---|---|
| 11. Quantitative variables | Explain how quantitative variables were handled in the analyses. If applicable, describe which groupings were chosen, and why. | Methods; Pages 7-10 |
| 12a. Statistical methods | Describe all statistical methods, including those used to control for confounding. | Methods; Page 10 |
| 12b. Statistical methods – subgroups and interactions | Describe any methods used to examine subgroups and interactions. | N/A |
| 12c. Statistical methods – missing data | Explain how missing data were addressed. | N/A |
| 12di. Statistical methods – loss to follow-up | **Cohort study:** If applicable, describe how loss to follow-up was addressed. | N/A |
| 12dii. Statistical methods – matching cases and controls | **Case-control study:** If applicable, explain how matching of cases and controls was addressed. | N/A |
| 12diii. Statistical methods – sampling strategy | **Cross-sectional study:** If applicable, describe analytical methods taking account of sampling strategy. | N/A |
| 12e. Statistical methods – sensitivity analyses | Describe any sensitivity analyses. | N/A |
| **Results** | | |
| 13a. Participant numbers | Report the numbers of individuals at each stage of the study—e.g., numbers potentially eligible, examined for eligibility, confirmed eligible, included in the study, completing follow-up, and analysed; Consider use of a flow diagram. | Results; Page 11 |
| 13b. Participants – non-participation | Give reasons for non-participation at each stage. | Results; Page 12 |
| 13c. Participants – flow diagram | Consider use of a flow diagram. | N/A |
| 14a. Descriptive data – participant characteristics | Give characteristics of study participants (e.g., demographic, clinical, social) and information on exposures and potential confounders. Present the information in a table. | Results; Page 11 |
| 14b. Descriptive data – missing data | Indicate the number of participants with missing data for each variable of interest. | Results; Page 12 |
| 14c. Descriptive data – follow-up time | **Cohort study:** Summarise follow-up time—e.g., average and total amount. | N/A |
| 15. Outcome data | **Cohort study:** Report numbers of outcome events or summary measures over time. **Case-control study:** Report numbers in each exposure category, or summary measures of exposure. **Cross-sectional study:** Report numbers of outcome events or summary measures. | N/A |

| | | |
|---|---|---|
| 16a. Main results | Give unadjusted estimates and, if applicable, confounder-adjusted estimates and their precision (e.g., 95% confidence intervals). Make clear which confounders were adjusted for and why they were included. | Results; Pages 11-15<br>Tables; Pages 25-28<br>Figures; Page 24 |
| 16b. Main results – category boundaries | Report category boundaries when continuous variables were categorised. | Results; Pages 11-15 |
| 16c. Main results – risk | If relevant, consider translating estimates of relative risk into absolute risk for a meaningful time period. | N/A |
| 17. Other analyses | Report other analyses done—e.g., analyses of subgroups and interactions, and sensitivity analyses. | N/A |
| **Discussion** | | |
| 18. Key results | Summarise key results with reference to study objectives. | Discussion; Page 15 |
| 19. Limitations | Discuss limitations of the study, taking into account sources of potential bias or imprecision. Discuss both direction and magnitude of any potential bias. | Discussion; Page 18 |
| 20. Interpretation | Give a cautious overall interpretation considering objectives, limitations, multiplicity of analyses, results from similar studies, and other relevant evidence. | Discussion; Pages 15-18 |
| 21. Generalisability | Discuss the generalisability (external validity) of the study results. | Discussion; Page 18 |
| **Other information** | | |
| 22. Funding | Give the source of funding and the role of the funders for the present study and, if applicable, for the original study on which the present article is based. | Study Funding; Page 20 |

### Appendix B – Exploratory Comprehensive Neuro-Ophtalmologic Assessment

Three patients (18.8% of our CAID cohort) underwent a comprehensive neuro-ophthalmologic evaluation. Slit lamp biomicroscopy, funduscopy, macular optical coherence tomography (OCT) and fluorescein retinal angiography were unremarkable in all cases. Ganglion cell layer and retinal nerve fiber layer OCT were normal in two patients, but showed mild bilateral and symmetrical atrophy in the third. Orthoptic assessment was normal in one patient; the second had comitant esotropia (likely due to decompensated esophoria) requiring prism correction; the third had minimal angle residual exotropia following a right medial rectus resection and a right lateral rectus recession procedure for intermittent exotropia during childhood (more than 20 years prior). Interestingly, the operative report stated that the extraocular muscles were poorly differentiated from the intermuscular septum. No other structural or vascular abnormalities were observed.

### Appendix C – Supplemental Figure

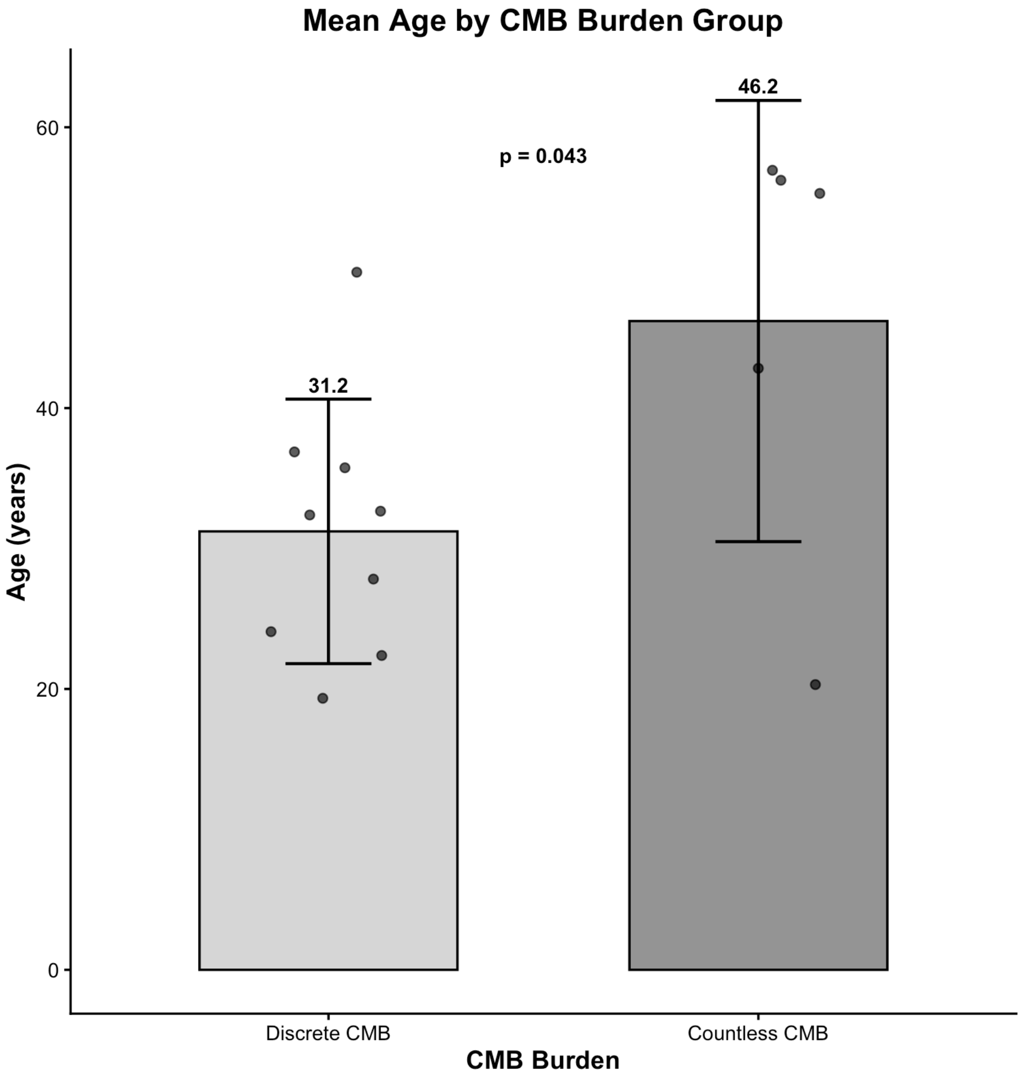


**Figure S1. Age comparison between cerebral microbleeds (CMB) burden groups.** Group differences were assessed using Student's t-test, with normality confirmed by the Shapiro–Wilk test.

**Appendix D – Local Cardiac Surgical Strategies in Patients with CAID Syndrome**

Several patients underwent cardiac surgery requiring cardiopulmonary bypass (CPB), including one for aortic valve replacement in the setting of bicuspid aortic valve and others during pacemaker implantation procedures, combined with atrial Maze surgery and left atrial appendage (LAA) exclusion.

LAA exclusion was performed using one of three approaches: percutaneous endovascular occlusion (plug device), surgical external clipping without CPB, or surgical ligation and excision under CPB. We preferentially performed surgical excision under CPB, as this technique is considered the most definitive and minimizes the risk of residual thrombogenic pouch formation, despite the need for extracorporeal circulation. No perioperative or postoperative intracranial hemorrhages were observed. Neurology consultation was systematically obtained preoperatively.

Our pacemaker implantation strategy evolved following recognition of the neurological phenotype associated with CAID syndrome. Transvenous systems are now avoided due to the risks of device-related infection and venous thrombosis or obstruction. Pacemakers are implanted via left thoracotomy using epicardial leads with a thoracic generator, typically positioned superior to the diaphragm. During the same procedure, LAA exclusion is performed either by surgical clipping (in patients considered at high hemorrhagic risk under CPB) or by CPB-assisted Maze procedure with LAA excision.

A limitation of this approach is the use of epicardial leads, which are not strictly MRI-conditional. However, the 2026 Canadian guidelines provide a more nuanced framework regarding MRI access in patients with non–MRI-conditional systems [1]. This may support future discussions with cardiology teams to facilitate broader access to cerebral MRI within this cohort.

**REFERENCE:**

1. Andrade JG, Joza J, Chew DS, et al. The Canadian Cardiovascular Society/Canadian Heart Rhythm Society Comprehensive Guidelines for the Selection, Implantation, and

Management of Patients With Cardiac Implantable Electronic Devices. Can J Cardiol 2026;42:1-68.
